# Hepatitis E in Bangladesh: Insights from a National Serosurvey

**DOI:** 10.1101/2021.06.14.21258872

**Authors:** Andrew S Azman, Kishor Kumar Paul, Taufiqur Rahman Bhuiyan, Aybüke Koyuncu, Henrik Salje, Firdausi Qadri, Emily S Gurley

**Affiliations:** Department of Epidemiology, Johns Hopkins Bloomberg School of Public Health, Baltimore, USA; icddrb, Dhaka, Bangladesh; University of Caimbridge, Cambridge, UK

**Keywords:** Hepatitis e, hepatitis E virus (HEV), seroprevalence

## Abstract

**Background:** Hepatitis E virus, typically genotypes 1 and 2, is a major cause of avoidable morbidity and mortality in South Asia. Although case fatality risk among pregnant women can reach as high as 25%, a lack of population-level disease burden data has been cited as a primary factor in key global policy recommendations against the routine use of licensed hepatitis E vaccines, one of the only effective tools available for preventing disease and death.

**Methods:** We tested serum from a nationally-representative serosurvey in Bangladesh for anti-HEV IgG. We estimated the proportion of the population with evidence of historical HEV infection and used Bayesian geostatistical models to generate high-resolution national maps of seropositivity. We examined variability in seropositivity by individual-level, household-level, and community-level risk factors using spatial logistic regression.

**Results:** We tested serum samples from 2924 individuals from 70 communities representing all divisions of Bangladesh and estimated a national seroprevalence of hepatitis E of 20% (95% CI 17-24%). Seropositivity increased with age and male sex (OR: 2.2, 95% CI: 1.8–2.8). Community-level seroprevalence ranged from 0-78% with the seroprevalence in urban areas being higher, including Dhaka, the capital, with 3-fold (95%CrI 2.3-3.7) higher seroprevalence than the rest of the country.

**Conclusion:** Hepatitis E infections are common throughout Bangladesh, though 90% of women reach reproductive age without any evidence of previous exposure to the virus, thus likely susceptible to infection and disease. Strengthening clinical surveillance for hepatitis E, especially in urban areas may help generate additional evidence needed to appropriately target interventions like vaccines to the populations most likely to benefit.

## Background

Hepatitis E is estimated to cause over 3 million cases of acute jaundice each year with more than 70,000 these leading to death and another 3000 still births [1]. Human infections from hepatitis E viruses, part of the orthohepevirus genus, are caused by four main genotypes (genotypes 1-4), with only genotypes 1 and 2 known to cause epidemics. HEV genotypes 1 and 2 are associated with self-limiting acute jaundice in the majority of infections although special populations, like pregnant women, have particularly poor outcomes with case fatality risk as high as 65% [2,3].

While HEV was only identified in 1981 [4], retrospective analyses have identified a number of large outbreaks which occurred on the Indian subcontinent in the 1970s and 1980s, including India and Bangladesh [5,6]. In Bangladesh, hepatitis E is endemic with large outbreaks from time-to-time [7–9]. Hepatitis E is the leading cause of acute jaundice in Bangladesh and may be responsible for up to 25% of maternal mortality [7,8,10].

While there is no effective treatment for acute hepatitis E and emergency improvements in water and sanitation have often been unsuccessful in curbing transmission [11], public health workers have very few effective tools at hand to mitigate the burden of outbreaks. Fortunately, a safe and efficacious vaccine is licensed in China and Pakistan and efforts are underway for licensure in other countries and WHO-prequalification [12]. A phase 4 clinical trial is on-going in Bangladesh [13], but no large-scale vaccination or other HEV-specific prevention efforts are planned, in-part due to our poor understanding of the burden and geographic distribution of the disease [14].

Following infection with hepatitis E, individuals develop long-lasting antibodies that can be measured through serosurveys to provide detailed insights into the history of infection in a population. Serosurveys can help us understand the geographic distribution and magnitude of historical HEV infections, identify risk factors and estimate key epidemiologic parameters related to transmission. Here we use a nationally representative serosurvey in Bangladesh to gain new insights into hepatitis E and provide critical details needed to target interventions, like vaccines, to areas at the highest risk.

## Methods

### Serosurvey Design

This survey was originally conducted as part of an arbovirus study in Bangladesh with two stage random sampling (community and household) previously described [15]. In brief, 70 communities from a total of 97,162 in the 2011 national census were selected with probability proportional to each community’s population. In rural areas (around three-quarters of the Bangladeshi population), the smallest administrative unit is a village, whereas in urban areas they are wards. Within each village or ward, study staff identified the household where community leaders said the most recent wedding had taken place and selected the nearest neighbor. From this neighboring household study staff chose a random direction and counted six households along a transect in that direction in order to identify the first potential study household. For subsequent households, study staff chose a random direction and selected the sixth household from the previous household in that community. In each selected household, study staff identified the household head, described the study, and invited them to participate in the study. If the household head agreed to participate, all household members older than 6 months of age were invited to take part. Within each community, study staff visited households until the day when at least 10 households had been visited with at least 40 serum samples. Within each household study staff administered structured questionnaires with questions about household-level infrastructure, wealth and assets in addition to individual data on demographics and travel history as well as collecting ∼5ml venous blood (∼3ml from children ≤3 years) from all consenting individuals. Data for this survey were collected from 10/2015 through 01/2016.

The study was approved by the icddr,b ethics review board (protocol number PR-14058); this secondary analysis was reviewed and deemed exempt from review by the Johns Hopkins Bloomberg School of Public Health Institutional Review Board. All adult participants provided written informed consent to participate in the study. Parents or guardians of all child participants provided written informed consent on their behalf.

### Laboratory Methods

Serum samples were stored at icddr,b at -80°C before testing. We tested the serum samples for the presence of anti-HEV IgG using the Wantai immunoassay kit (Wantai HEV IgG [WE-7296] ELISA kit; Wantai Biological, Beijing, China) following manufacturer’s instructions. As suggested in the packet insert, samples with a standardized optical density > 1.1 were considered positive, those < 0.9 were considered negative, and those in the range 0.9–1.1 were considered indeterminate.

### Statistical Analyses

We estimated the national seroprevalence by including survey design weights and post-stratifying by age and sex to the 2011 Bangladesh census, with confidence intervals estimated using the Rao-Scott method implemented in the survey package for R [16,17]. We used the same approach to estimating seroprevalence by urban/rural locations, sex and age (only post-stratifying by sex for age-group-specific estimates). We excluded indeterminate results from all primary analyses.

We explored the relationship between individual, household and community-level factors and seropositivity using hierarchical logistic regression models including a spatial random field assuming a Matern covariance structure using integrated nested Laplace approximations (INLA) as implemented in the R-INLA package [18]. All individual and household-level data were collected from the survey questionnaire. Community-level data for population density [19], travel time to the nearest city [20], distance to a major water body, altitude, and poverty index [21] were collected from publicly available data sources. In the main analyses we included household and community random effects in addition to the spatial random field, but in sensitivity analyses estimated models with different combinations of random effects to understand variability in our estimates (Supplement). We explored univariate models, a fully saturated model, a model with only variables significant in the univariate analyses and two simplified models selected *a priori* and compared their fit with Wanatabe-Akaike information criterion (WAIC) [22].

Using the same INLA modeling framework we estimated seroprevalence on a 5 km by 5 km grid across Bangladesh. To do this we assigned each community to a grid-cell by its centroid, estimated the mean seroprevalence in each cell containing observations and fit spatial regression models to these data. We then used these fitted models to predict seroprevalence in the unobserved grid-cells. We fit both a fully saturated model, including population size, distance from a major water body, a poverty index, travel time to the nearest city, and altitude as linear predictors in addition to spatial random effects using a Matern covariance structure and a null model with only the spatial random effects. To quantify the out of sample performance of this approach we used leave-one-community out cross validation and compared predictions to a ‘naive’ model that predicted the mean grid-cell seroprevalence for all but the held-out cells and calculated the mean absolute error.

We predicted seropositivity among girls reaching child-bearing age (15 years) by fitting generalized additive models with penalized cubic splines to age-seroprevalence curves in each first-level administrative unit (division). We estimated simultaneous 95% confidence intervals by re-sampling from estimates of the variance-covariance matrix of the fitted model using a simulation-based approach with 1,000 draws [23].

We used the GADM 3.6 spatial database for all administrative boundaries, which does not include boundary changes made after September, 2015. All analyses were performed in R (version 4.0.2). Data and source code to reproduce analyses are available at https://github.com/HopkinsIDD/hepE-bangladesh-national-serosurvey.

## Results

We tested 2924 individuals from 707 households and 70 communities representing all first-level administrative units (divisions) of Bangladesh. Sampled individuals had a similar age and sex distribution to the population of Bangladesh with the exception of young children, who were under-represented [24].

Overall, 20.9% (610) of individuals tested positive for anti-HEV IgG, 78.8% (2305) were negative and 0.3% (9) were indeterminate. After taking into account the survey design and adjusting for imbalances between the sampled population and that of Bangladesh, we estimated a national seroprevalence of 20.0% (95% Confidence Interval [CI] 16.5-23.9%; design effect = 6.4).

Seroprevalence increased with age, from 2.5% (95%CI 0.6-9.7) in those under 5 years old and reaching a maximum of 40.9% (95%CI 26.0-57.8) among those 70-74 years old. On average, men (24.3%; 95%CI 20.5-28.5) had higher seroprevalence than women (15.9%; 95%CI 12.4-20.3) with this pattern holding across most age groups (Figure 1). Nationally, 90.4% (95%CI 88.4-92.1%) of girls reach the age of 15 (roughly “reproductive age”) without evidence of antibodies and likely susceptible to disease [25].

**Figure 1.**
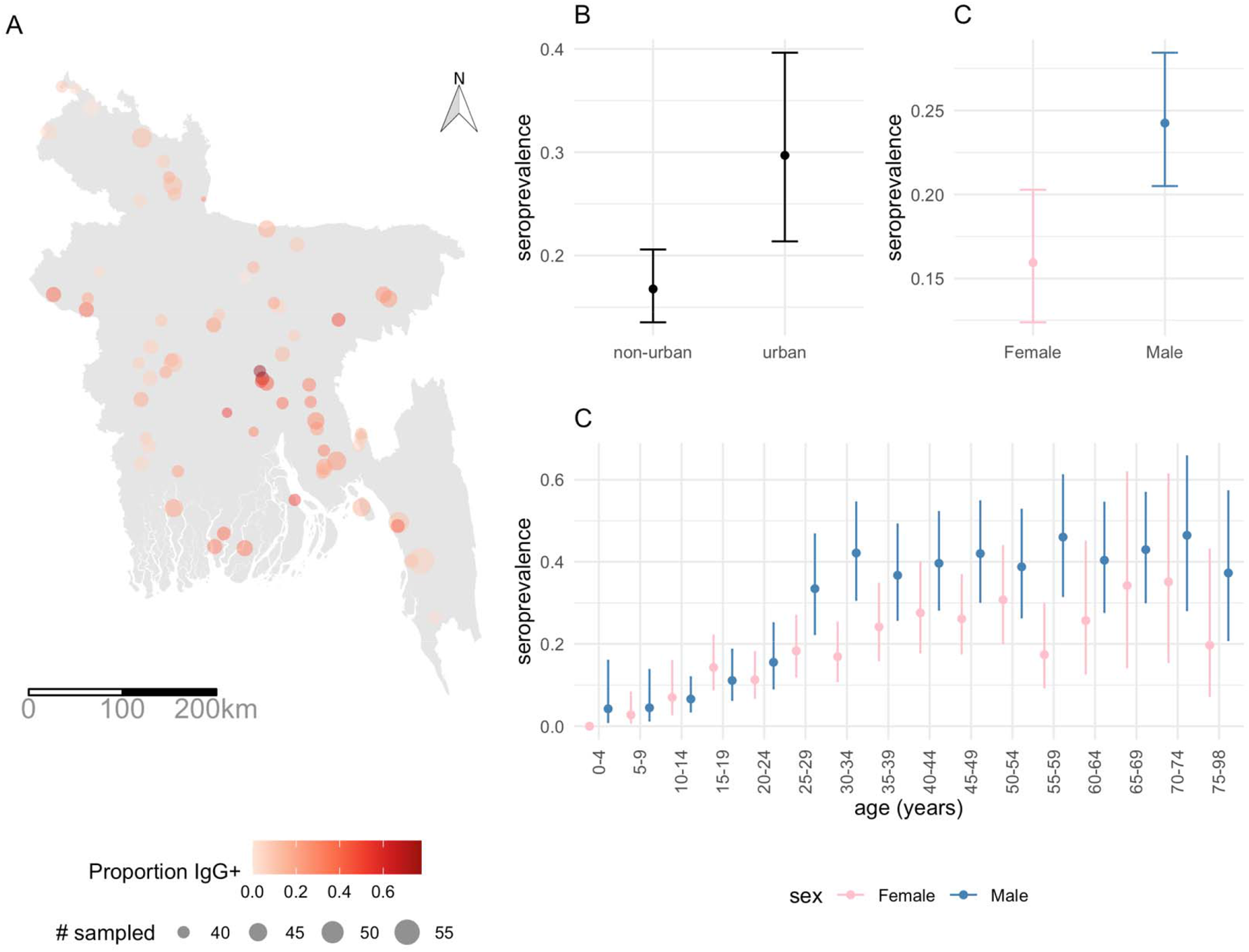
Overview of sampled individuals and communities and seroprevalence estimates. Panel A illustrates the sampled community locations, number of individuals sampled (size of dots) and the proportion of individuals seropositive (color). Panel B illustrates the adjusted seroprevalence (age/sex) by urban/non-urban classifications and Panels C and D illustrate the sex (adjusted for age) and age/sex-specific seroprevalence.

Seroprevalence varied greatly across communities from 0% to 77.5% across the country (Figure 1). Urban areas (29.7%; 95%CI 21.4-39.6) had 1.8 times higher seroprevalence than rural areas (16.8%; 95%CI 13.5-20.6). Household-level seroprevalence ranged from 0 to 100% and we found no evidence of increasing household seroprevalence with household size.

While age, sex and living in an urban area were associated with the risk of being seropositive, other community and household-level risk factors may also be important. To explore the relationship between these potential risk factors we used a series of univariable and multivariable logistic regression models with spatially correlated errors. In univariate models (Table 1), we found significant positive associations with age, being male, travel, urbanicity, population density, a community poverty index, and protective effects of various indicators of socioeconomic status (e.g., having completed primary school compared to having no formal education, having cattle or other animals in the household, being a household owner). However, in our primary multivariable model with spatial random effects, none of these factors were independently associated with seropositivity except for age and sex, though the effect sizes were largely consistent across various models considered (Table 1 and Supplementary Table 1). Estimates from models with varying assumptions about random effects yielded qualitatively similar results.

**Table 1.**
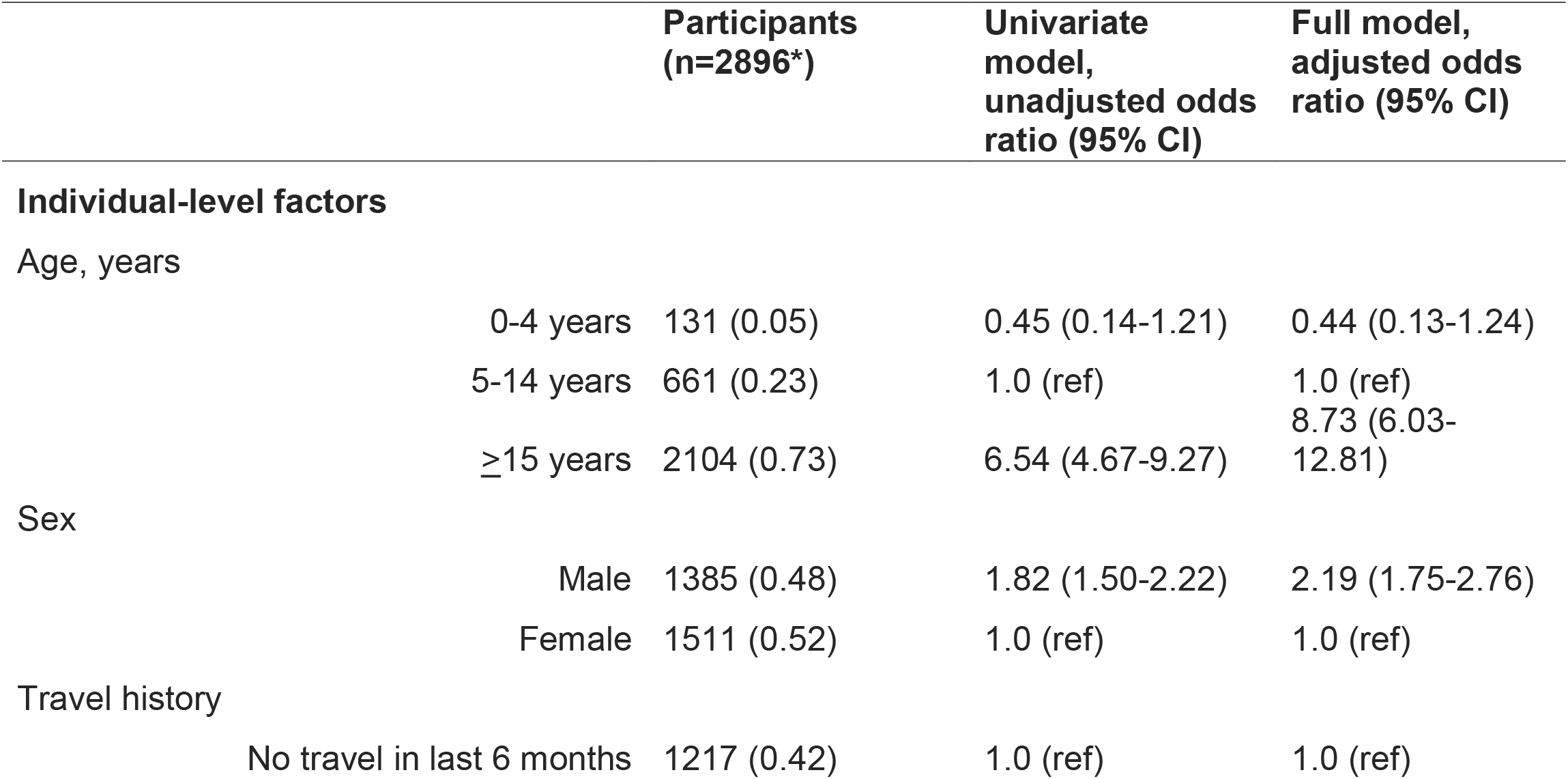
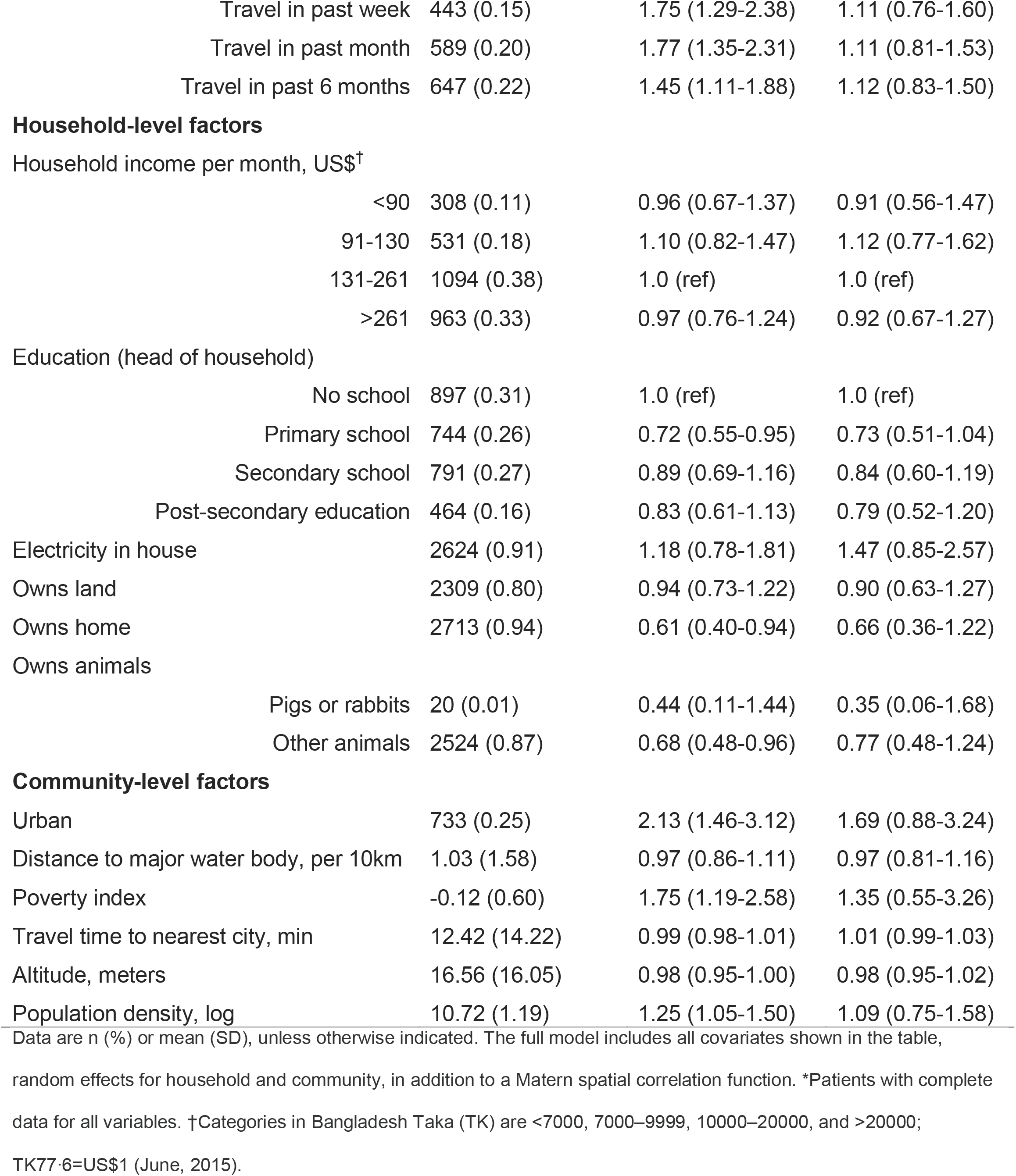
Estimated odds ratios and 95% credible intervals for seropositivity including random effects for household, community and a spatial random field.

|  |  | Participants<br>(n=2896*) | Univariate<br>model,<br>unadjusted odds<br>ratio (95% CI) | Full model,<br>adjusted odds<br>ratio (95% CI) |
| --- | --- | --- | --- | --- |
| **Individual-level factors** |  |  |  |  |
| Age, years |  |  |  |  |
|  | 0-4 years | 131 (0.05) | 0.45 (0.14-1.21) | 0.44 (0.13-1.24) |
|  | 5-14 years | 661 (0.23) | 1.0 (ref) | 1.0 (ref) |
|  | ≥15 years | 2104 (0.73) | 6.54 (4.67-9.27) | 8.73 (6.03-12.81) |
| Sex |  |  |  |  |
|  | Male | 1385 (0.48) | 1.82 (1.50-2.22) | 2.19 (1.75-2.76) |
|  | Female | 1511 (0.52) | 1.0 (ref) | 1.0 (ref) |
| Travel history |  |  |  |  |
|  | No travel in last 6 months | 1217 (0.42) | 1.0 (ref) | 1.0 (ref) |

|  |  |  |  |
| --- | --- | --- | --- |
| Travel in past week | 443 (0.15) | 1.75 (1.29-2.38) | 1.11 (0.76-1.60) |
| Travel in past month | 589 (0.20) | 1.77 (1.35-2.31) | 1.11 (0.81-1.53) |
| Travel in past 6 months | 647 (0.22) | 1.45 (1.11-1.88) | 1.12 (0.83-1.50) |

|  |  |  |  |
| --- | --- | --- | --- |
| <90 | 308 (0.11) | 0.96 (0.67-1.37) | 0.91 (0.56-1.47) |
| 91-130 | 531 (0.18) | 1.10 (0.82-1.47) | 1.12 (0.77-1.62) |
| 131-261 | 1094 (0.38) | 1.0 (ref) | 1.0 (ref) |
| >261 | 963 (0.33) | 0.97 (0.76-1.24) | 0.92 (0.67-1.27) |

Education (head of household)
|  |  |  |  |
| --- | --- | --- | --- |
| No school | 897 (0.31) | 1.0 (ref) | 1.0 (ref) |
| Primary school | 744 (0.26) | 0.72 (0.55-0.95) | 0.73 (0.51-1.04) |
| Secondary school | 791 (0.27) | 0.89 (0.69-1.16) | 0.84 (0.60-1.19) |
| Post-secondary education | 464 (0.16) | 0.83 (0.61-1.13) | 0.79 (0.52-1.20) |

|  |  |  |  |
| --- | --- | --- | --- |
| Pigs or rabbits | 20 (0.01) | 0.44 (0.11-1.44) | 0.35 (0.06-1.68) |
| Other animals | 2524 (0.87) | 0.68 (0.48-0.96) | 0.77 (0.48-1.24) |

We fitted Bayesian geostatistical models to make a national map of seroprevalence. Our primary model demonstrated out-of-sample predictive skill (mean absolute error = 10.6%) with little bias (1.85 × 10^−4^) and moderate correlation of predictions with the true values in cross-validation (Pearson’s correlation) of 0.51 (Figure 2). The seroprevalence map reveals large heterogeneity in seroprevalence across the country with the highest seroprevalence around Dhaka with some evidence of higher-than-average risk in two other large cities, Chittagong and Rajshahi. Similar to the non-spatial analyses, we estimate from these maps that 21.6% (95% CI 19.0-24.3%) of the population has been infected during their lifetime (35,177,057 people, 95%CrI 30,919,165-39,527,957). Residents of Dhaka have a 3.0-fold (95%CrI 2.3-3.7) higher seroprevalence than the mean seroprevalence of the rest of Bangladesh. Alternative models including different combinations of random effects (household, community and spatial) and covariates led to similar maps.

**Figure 2.**
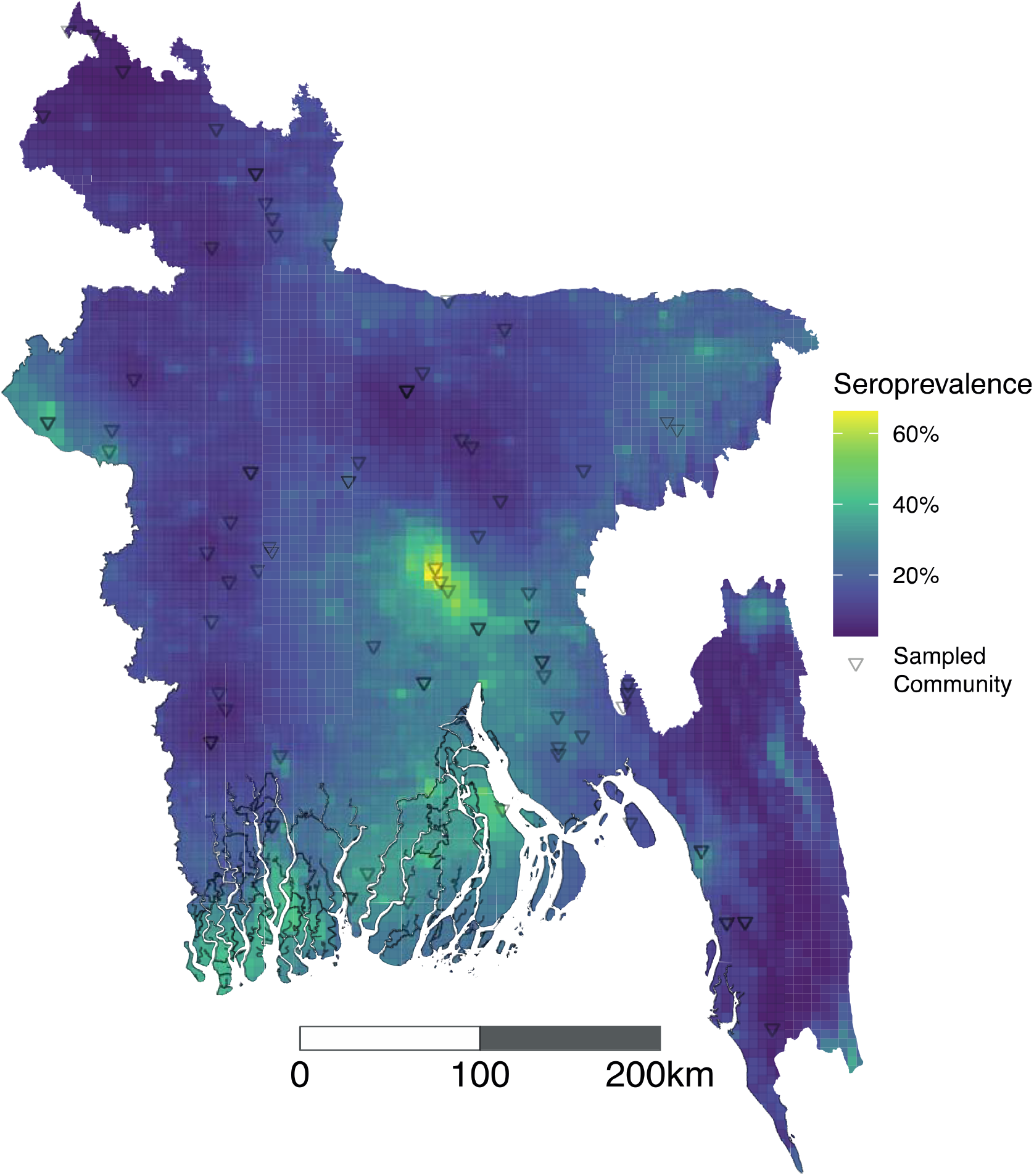
Predicted seroprevalence across Bangladesh, 2015. Predicted seroprevalence from geostatistical model with distance from major water body, population density, altitude, poverty index, and travel time to nearest city.

## Discussion

In this nationally representative serosurvey we found that one in five people in Bangladesh had evidence of prior HEV infection with men having more than 1.5 times higher risk than women. Seroprevalence was 3 times higher in Dhaka, Bangladesh’s capital and largest city than the rest of the country. Given the lack of specificity of clinical case definitions for hepatitis E (i.e., acute jaundice syndrome) and limited laboratory diagnostic use across Bangladesh, our approach and results highlight an important avenue for understanding risk across the country with an aim of targeting surveillance, prevention and control activities.

Although faecal contamination of drinking water is likely the predominant cause of HEV infections in LMICs [26], measures of socioeconomic status typically associated with household access to clean water and sanitation (e.g. household income, education level) were not significantly predictive of seropositivity in our study. Our data are from a study not originally designed to study hepatitis E, therefore, we did not have data on household water sources and sanitation. Despite this, our findings are consistent with existing literature on risk factors for HEV infection in Bangladesh [27,28] and documented large outbreaks of hepatitis E associated with contaminated municipal water supplies in urban areas of Bangladesh [8]. The higher seroprevalence among males may be due to exposures outside of the home, given their propensity to leave home more often than women in Bangladesh. If this hypothesis is correct, household-level water and sanitation interventions alone may not be sufficient to interrupt HEV transmission. Water and sanitation interventions may also have limited utility in preventing sporadic acute hepatitis cases associated with exposure to blood and animals, which are hypothesized to contribute to the burden of hepatitis E in Bangladesh [29] and other LMICs [30].

Samples from this same serosurvey were previously used to map the annual risk of *Vibrio cholerae* O1 infections across the country [31]. While both *Vibrio cholerae* and hepatitis E are transmitted through fecal contamination of drinking water and food, the spatial distribution of risk of these infections are completely different in Bangladesh. For example, while many *V. cholerae* infections were estimated to occur in Dhaka, inhabitants had lower than average risk overall. In contrast, inhabitants of Dhaka had 3 times higher risk of HEV seropositivity than others in the country. Some of the differences in spatial risk profiles may be due to the fact that cholera estimates capture only a snapshot of transmission (one year) compared to the lifetime exposures captured by HEV antibodies. Additionally, men had significantly higher HEV seroperelance though we found no significant difference by sex for cholera. These differences might be in part explained by HEV transmission being facilitated through urban water infrastructure (e.g., [8]) and cholera transmission occurring more broadly through fecal oral routes inside households [32].

This study comes with a number of limitations. We assumed that these serologic assays had perfect sensitivity and specificity for detecting historical HEV infections. Previous studies have estimated sensitivity and specificity of these assays to be high [33,34] however, without a gold standard assay to compare against these estimates are unlikely to be perfect nor generalisable to all settings. Furthermore, seropositivity has been shown to decay over time so those infected many years before the serosurvey may be differentially misclassified as seronegative [35,36]. That seroprevalence does not significantly increase past the age of ∼30 years old is likely due to a combination of seroreversion and changes in the force of infection over the decades. Future work synthesizing assay validation data may be valuable for correcting seroprevalence estimates appropriately. As there is only one HEV serotype, our estimates likely include immunologically meaningful exposures not only to genotypes 1 and 2, the most concerning for outbreaks, but other genotypes, which have not been widely described in Bangladesh and lead to different, though still severe, clinical outcomes. While we present smoothed estimates of the seroprevalence throughout the country, these are based on a geostatistical model fit to data from only 70 sampled communities throughout the country. These models assume that risk varies smoothly across space after taking into account covariates though true HEV risk is likely less smooth over space. Finally, the household sampling approach used to recruit individuals in the parent study may have systematically excluded migrant populations and individuals living in informal settlements more likely to have inadequate WASH and more likely to be seropositive.

Our estimates provide a snapshot of cumulative infection risk in Bangladesh in 2015/2016. While this is useful to understand large-scale differences in risk across the country, it masks important differences in risk over time and space. The age-stratified patterns of seroprevalence, and in particular the changes in seroprevalence among the youngest children can be particularly informative for understanding recent infection risk (i.e., force of infection), which may be more important for guiding policy. Our sample size in each sampled village of children under 5 years old was too small to permit detailed age-stratified analysis in these young age groups though future serosurveys may benefit greatly from these individuals. Furthermore, longitudinal or repeated cross-sectional serosurveys can allow for estimates of seroincidence [28]. Estimates of the contemporary force of infection can be combined with data on the proportion of infections that become clinically apparent (and severe) to help estimate the burden of hepatitis E [1].

Given that more than 90% of girls reaching child-bearing age are susceptible to infection and the high case fatality risk associated with hepatitis E in pregnant women [8,26], this group may benefit greatly from hepatitis E vaccine [37] and existing maternal and child health programs in LMICs such as Bangladesh may be a cost-effective platform to facilitate delivery. The World Health Organization has not recommended routine use of this vaccine due to a lack of data on the safety, immunogenicity and efficacy of the vaccine among pregnant women as well as a lack of epidemiologic data on the burden of disease in the general population [12]. Fortunately, a clinical trial evaluating the safety, immunogenicity and effectiveness of hepatitis E vaccines among women of childbearing age, including those that go on to become pregnant, are underway [13] in Bangladesh. Data from our study suggest that these vulnerable individuals are at high exposure risk across the country and the use of safe and effective hepatitis E vaccines among pregnant women in Bangladesh may be justified.

In this study we illustrate how remnant samples from population-based serologic studies not originally obtained to study hepatitis E can be an effective strategy to generate critical epidemiologic data in LMICs where surveillance infrastructure is weak or nonexistent. The SARS-CoV-2 pandemic presents an unprecedented opportunity to leverage the increased number of representative population-based surveys to improve our understanding of the global burden of hepatitis E [38,39]. Countries currently planning serial cross-sectional serosurveys to monitor trends in SARS-CoV-2 transmission [40] should consider utilizing remnant samples to generate data that may help quantify hepatitis E risk over time and accelerate the use of the licensed vaccine.

## Data Availability

Data and source code to reproduce analyses are available at https://github.com/HopkinsIDD/hepE-bangladesh-national-serosurvey.

## Funding statement

The original data collection was supported by the Centers for Disease Control and Prevention (5U01GH001207-02). icddr,b acknowledges the commitment of the US CDC to its research efforts and thanks the Governments of Bangladesh, Canada, Sweden, and the UK for providing core or unrestricted support. US National Institutes for Health (NIH; R01 AI135115 ASA). Support to FQ and her laboratory also came from the NIH Fogarty International Center (TW005572) and Emerging Global Leader Award (K43 TW010362). The funders had no role in study design, data collection and interpretation, or the decision to submit the work for publication.

## Acknowledgement

We thank all the participants of the study throughout Bangladesh for their willingness to participate in this research. We also wish to thanks the field and laboratory staff for their dedicated work.

## Conflicts of interest

The authors have no conflicts of interest to declare.

## Notes

### Competing Interest Statement

The authors have declared no competing interest.

## References

1. Rein DB, Stevens GA, Theaker J, Wittenborn JS, Wiersma ST. The global burden of hepatitis E virus genotypes 1 and 2 in 2005. Hepatology. Wiley; 2012; 55(4):988–997.

2. Kamar N, Bendall R, Legrand-Abravanel F, et al. Hepatitis E. Lancet. 2012; 379(9835):2477– 2488.

3. Amanya G, Kizito S, Nabukenya I, et al. Risk factors, person, place and time characteristics associated with Hepatitis E Virus outbreak in Napak District, Uganda. BMC Infect Dis. 2017; 17(1):451.

4. Balayan MS, Andjaparidze AG, Savinskaya SS, et al. Evidence for a virus in non-A, non-B hepatitis transmitted via the fecal-oral route. Intervirology. 1983; 20(1):23–31.

5. Khuroo MS. Study of an epidemic of non-A, non-B hepatitis. Possibility of another human hepatitis virus distinct from post-transfusion non-A, non-B type. Am J Med. 1980; 68(6):818–824.

6. Hlady WG, Islam MN, Wahab MA, Johnson SD, Waiz A, Krawczynski KZ. Enterically transmitted non-A, non-B hepatitis associated with an outbreak in Dhaka: epidemiology and public health implications. Trop Doct. 1990; 20(1):15–17.

7. Paul RC, Nazneen A, Banik KC, et al. Hepatitis E as a cause of adult hospitalization in Bangladesh: Results from an acute jaundice surveillance study in six tertiary hospitals, 2014-2017. PLoS Negl Trop Dis. 2020; 14(1):e0007586.

8. Gurley ES, Hossain MJ, Paul RC, et al. Outbreak of hepatitis E in urban Bangladesh resulting in maternal and perinatal mortality. Clin Infect Dis. 2014; 59(5):658–665.

9. Biswas RSR, Sydu S, Mamun SMH, et al. Viral load and genotype of recent hepatitis E virus outbreak in Chittagong, Bangladesh. Journal of the Scientific Society. Medknow Publications and Media Pvt. Ltd.; 2020; 47(3):144.

10. Gurley ES, Halder AK, Streatfield PK, et al. Estimating the burden of maternal and neonatal deaths associated with jaundice in Bangladesh: possible role of hepatitis E infection. Am J Public Health. 2012; 102(12):2248–2254.

11. Guthmann J-P, Klovstad H, Boccia D, et al. A large outbreak of hepatitis E among a displaced population in Darfur, Sudan, 2004: the role of water treatment methods. Clin Infect Dis. 2006; 42(12):1685–1691.

12. World Health Organization. Recommendations to assure the quality, safety and efficacy of recombinant hepatitis E vaccines [Internet]. 2018. Available from: https://www.who.int/biologicals/expert_committee/POST_ECBS_2018_Recommendations_HEP_E_vaccines.pdf

13. Zaman K, Dudman S, Stene-Johansen K, et al. HEV study protocol : design of a cluster-randomised, blinded trial to assess the safety, immunogenicity and effectiveness of the hepatitis E vaccine HEV 239 (Hecolin) in women of childbearing age in rural Bangladesh. BMJ Open. 2020; 10(1):e033702.

14. WHO. Hepatitis E vaccine: WHO position paper, May 2015--Recommendations. Vaccine. 2016; 34(3):304–305.

15. Salje H, Paul KK, Paul R, et al. Nationally-representative serostudy of dengue in Bangladesh allows generalizable disease burden estimates. Elife [Internet]. 2019; 8. Available from: http://dx.doi.org/10.7554/eLife.42869

16. Rao JNK, Scott AJ. On Chi-Squared Tests for Multiway Contingency Tables with Cell Proportions Estimated from Survey Data [Internet]. The Annals of Statistics. 1984. Available from: http://dx.doi.org/10.1214/aos/1176346391

17. Lumley T. Complex Surveys: A Guide to Analysis Using R. John Wiley & Sons; 2011.

18. Lindgren F, Rue H, Lindström J. An explicit link between Gaussian fields and Gaussian Markov random fields: the stochastic partial differential equation approach. J R Stat Soc Series B Stat Methodol. Wiley; 2011; 73(4):423–498.

19. Tatem AJ. WorldPop, open data for spatial demography. Sci Data. 2017; 4:170004.

20. Weiss DJ, Nelson A, Gibson HS, et al. A global map of travel time to cities to assess inequalities in accessibility in 2015. Nature. 2018; 553(7688):333–336.

21. Steele JE, Sundsøy PR, Pezzulo C, et al. Mapping poverty using mobile phone and satellite data. J R Soc Interface [Internet]. 2017; 14(127). Available from: http://dx.doi.org/10.1098/rsif.2016.0690

22. Watanabe K. An alternative view of variational Bayes and asymptotic approximations of free energy. Mach Learn. 2012; 86(2):273–293.

23. Ruppert D, Wand MP, Carroll RJ. Semiparametric Regression. Cambridge University Press; 2003.

24. Azman AS, Lauer S, Bhuiyan MTR, et al. Vibrio cholerae O1 transmission in Bangladesh: insights from a nationally-representative serosurvey. medRxiv [Internet]. Cold Spring Harbor Laboratory Press; 2020;. Available from: https://www.medrxiv.org/content/10.1101/2020.03.13.20035352v1.abstract

25. Choi Y, Zhang X, Skinner B. Analysis of IgG Anti-HEV Antibody Protective Levels During Hepatitis E Virus Reinfection in Experimentally Infected Rhesus Macaques. J Infect Dis. 2019; 219(6):916–924.

26. Organization WH, Others. The global prevalence of hepatitis A virus infection and susceptibility: a systematic review. https://apps.who.int › iris › handle https://apps.who.int › iris › handle [Internet]. World Health Organization; 2010;. Available from: https://apps.who.int/iris/handle/10665/70513

27. Labrique AB, Zaman K, Hossain Z, et al. Population seroprevalence of hepatitis E virus antibodies in rural Bangladesh. Am J Trop Med Hyg. 2009; 81(5):875–881.

28. Labrique AB, Zaman K, Hossain Z, et al. Epidemiology and risk factors of incident hepatitis E virus infections in rural Bangladesh. Am J Epidemiol. 2010; 172(8):952–961.

29. Sazzad HMS, Luby SP, Labrique AB, et al. Risk Factors Associated with Blood Exposure for Sporadic Hepatitis E in Dhaka, Bangladesh. Am J Trop Med Hyg. 2017; 97(5):1437–1444.

30. Koyuncu A, Mapemba D, Ciglenecki I, Gurley ES, Azman AS. Setting a course for preventing hepatitis E in low and lower-middle-income countries: A systematic review of burden and risk factors. Open Forum Infect Dis [Internet]. Oxford University Press; 2021 [cited 2021 May 14];. Available from: https://academic.oup.com/ofid/advance-article/doi/10.1093/ofid/ofab178/6224343

31. Azman AS, Lauer SA, Bhuiyan TR, et al. Vibrio cholerae O1 transmission in Bangladesh: insights from a nationally representative serosurvey. Lancet Microbe. 2020; 1(8):e336–e343.

32. Azman AS, Luquero FJ, Salje H, et al. Micro-Hotspots of Risk in Urban Cholera Epidemics. J Infect Dis. 2018; 218(7):1164–1168.

33. Avellon A, Morago L, Garcia-Galera del Carmen M, Munoz M, Echevarría J-M. Comparative sensitivity of commercial tests for hepatitis E genotype 3 virus antibody detection. J Med Virol. 2015; 87(11):1934–1939.

34. Kodani M, Kamili NA, Tejada-Strop A, et al. Variability in the performance characteristics of IgG anti-HEV assays and its impact on reliability of seroprevalence rates of hepatitis E. J Med Virol. 2017; 89(6):1055–1061.

35. Kmush BL, Zaman K, Yunus M, Saha P, Nelson KE, Labrique AB. A Ten Year Immunopersistence Study of Hepatitis E Antibodies in Rural Bangladesh. Am J Epidemiol [Internet]. 2018;. Available from: http://dx.doi.org/10.1093/aje/kwy044

36. Faber M, Willrich N, Schemmerer M, et al. Hepatitis E virus seroprevalence, seroincidence and seroreversion in the German adult population. J Viral Hepat. 2018; 25(6):752–758.

37. Zhu F-C, Zhang J, Zhang X-F, et al. Efficacy and safety of a recombinant hepatitis E vaccine in healthy adults: a large-scale, randomised, double-blind placebo-controlled, phase 3 trial. Lancet. 2010; 376(9744):895–902.

38. Chen X, Chen Z, Azman AS, et al. Serological evidence of human infection with SARS-CoV-2: a systematic review and meta-analysis. The Lancet Global Health. Elsevier; 2021; 9(5):e598–e609.

39. Mina MJ, Metcalf CJE, McDermott AB, Douek DC, Farrar J, Grenfell BT. A Global lmmunological Observatory to meet a time of pandemics. Elife. eLife Sciences Publications, Ltd; 2020; 9:e58989.

40. Kumar MS, Bhatnagar T, Manickam P, et al. National sero-surveillance to monitor the trend of SARS-CoV-2 infection transmission in India: Protocol for community-based surveillance. Indian J Med Res. 2020; 151(5):419–423.

